# Laboratory evaluation of five rapid diagnostic tests for the serological diagnosis of chronic *Trypanosoma cruzi* infection in Yucatán, Mexico

**DOI:** 10.64898/2026.10.26.26366472

**Authors:** R Heslop, G Pellegrin, PP Martinez-Vega, CF Teh-Poot, J Torres-Montero, H Noh-Pech, KR Dzul-Rosado, F Puerto-Manzano, A Gonzalez-Martínez, CN Ibarra-Cerdeña, CG Meza-Ayala, O Valenzuela-Antelo, JV Cruz-Chan, V Lejon, C Guzman-Bracho, JA Ortiz, A Ramos-Ligonio, J Alonso-Padilla, B Bucheton, E Waleckx

## Abstract

Chagas disease (CD), caused by *Trypanosoma cruzi*, is a significant public health challenge in Latin America, where an estimated 70% of infected individuals remain undiagnosed. Rapid diagnostic tests (RDTs) offer a promising solution for new screening strategies, yet their performance has not been systematically assessed in Mexican cohorts.

Here, we evaluated the diagnostic performance of five RDTs: Chagas Rapid Test (Certum), Bio-Chagas Rapid Test (Mexlab), Bioline^TM^ Chagas Ab (Abbott), Chagas Rapid Test (Amunet) and Chagas Sero *K*-SeT (Coris BioConcept). Performance was tested using 79 archived human serum samples from Yucatán. All tests were benchmarked against the diagnostic reference algorithm established by the Mexican Official Norm (*NOM*), where samples were characterised as positive if they were reactive to ≥ 2 conventional serological assays (Chagatest *Lisado*, Chagatest *Recombinante* v4.0 and Chagatest HAI).

The RDTs demonstrated variable diagnostic sensitivity. The Bioline^TM^ Chagas Ab test demonstrated the highest sensitivity (97.2%, 95% CI: 85.5 - 99.9; 35/36 positive samples detected) and almost perfect statistical agreement with the *NOM* reference algorithm (κ= 0.90; *p* < 0.0001). Similarly, the Chagas Sero *K*-SeT prototype showed a high sensitivity (91.7%, 95% CI: 77.5 – 98.2; 33/36 positive samples detected) and substantial concordance (κ= 0.87, *p* < 0.0001) with the *NOM*-based composite result. In contrast, the remaining RDTs demonstrated low sensitivities (Certum: 0.0%, 95% CI: 0.0–9.7; Mexlab: 5.6%, 95% CI: 0.7–18.7; Amunet: 33.3%, 95% CI: 18.6–51.0), limiting their suitability for CD diagnostic algorithms, at least in Yucatán. Specificity was consistently high across the RDTs, ranging from 93.0% to 100% (95% CIs: 80.9–100). These findings underscore the need to locally validate diagnostic tools. The Bioline^TM^ Chagas Ab and Chagas Sero *K*-SeT tests met the WHO benchmarks for RDT performance and, if validated in larger cohorts, could strengthen the screening of *T. cruzi* infection and epidemiological surveillance in Mexico.

**Author summary:** Chagas disease, caused by the parasite *Trypanosoma cruzi*, remains underdiagnosed in Mexico. Rapid diagnostic tests (RDTs) could help expand access to screening, particularly in settings where laboratory testing is difficult to implement. However, the performance of these tests can vary geographically and has not been systematically evaluated in Mexico.

We evaluated four commercially available RDTs and one developmental prototype using archived serum samples from Yucatán, a region where Chagas disease is endemic. Results were compared with the diagnostic algorithm recommended by the Mexican Official Norm.

The Bioline™ Chagas Ab test showed the highest sensitivity, detecting 35 of 36 positive samples, while the Chagas Sero K-SeT prototype detected 33. The other three commercial RDTs showed substantially lower sensitivity, detecting between 0 and 12 positive samples. Specificity was high across all five tests, ranging from 93.0% to 100%, indicating that most negative samples were correctly identified. However, the low sensitivity of three tests limits their usefulness for Chagas disease screening in Yucatán.

Our findings highlight the importance of locally validating RDTs before their implementation. The Bioline^TM^ Chagas Ab and Chagas Sero *K*-SeT tests show promise for Chagas disease screening in Mexico but require further evaluation using larger and geographically diverse cohorts.

## Background

Chagas disease (CD) is a neglected tropical disease caused by the protozoan parasite *Trypanosoma cruzi* (WHO, 2025). It is mainly transmitted to humans and other mammals through the contaminated faeces of hematophagous triatomine bugs (family Reduviidae), either through abrasions in the skin, mucous membranes or ingestion of contaminated food or drinks (de Sousa *et al*., 2023). However, non-vectorial routes, such as blood transfusion, organ transplantation and congenital transmission, also contribute to the disease burden.

CD is endemic in 21 Latin American countries, where it is estimated that more than 7.4 million people are infected with *T. cruzi*, and more than 104 million people are at risk of infection (PAHO, 2025). CD causes over 10,000 deaths every year. However, systemic barriers to routine screening and confirmatory diagnosis prevent approximately 70% of infected individuals from receiving a diagnosis (Cucunubá et al., 2024; Schijman et al., 2024). Furthermore, less than 10% of those diagnosed will receive antiparasitic treatment with either benznidazole or nifurtimox, the two available drugs against *T. cruzi* (PAHO, 2022).

Mexico is endemic for Chagas disease (CD), where the highest numbers of reported cases occur in the southern states of Yucatán, Veracruz, Chiapas, Quintana Roo, and Oaxaca, as well as in the central state of Morelos (Ibáñez-Cervantes *et al*., 2018). National seroprevalence has previously been estimated at 3.38%, corresponding to approximately 4.06 million infections (Arnal *et al.,* 2019). Current diagnostic algorithms to detect *T. cruzi* infection are performed according to the Official Mexican Standard (*NOM*, *Norma Oficial Mexicana*; ref.: *NOM-032-SSA2-2014*). In the acute phase, diagnosis relies on identifying epidemiological exposure to the vector and compatible inflammatory signs and is confirmed by detecting *T. cruzi* through direct parasitological methods (e.g., thick or thin blood smear microscopy, microhematocrit, blood culture) or PCR. Evidence of seroconversion by Enzyme-Linked Immunosorbent Assay (ELISA), indirect immunofluorescence (IIF), or haemagglutination inhibition (HAI) after four weeks of infection supports the diagnosis. However, the acute phase is often asymptomatic (∼95% cases) and is rarely diagnosed (Rassi et al., 2010). The majority of cases are diagnosed exclusively by serology during the chronic stage of the infection (Schijman et al., 2024). A positive diagnosis requires reactive results from two or more distinct serological tests utilising different antigen targets (*NOM-032-SSA2-2014*). However, previous studies have demonstrated that conventional serological test performance in Mexico may be lower than that reported in other endemic regions (Truyens et al. 2021). Although the *NOM* algorithm provides clear guidelines, few cases of *T. cruzi* infection in Mexico are diagnosed via clinical suspicion and evaluation (Aké-Chan et al, 2024). Instead, incidental diagnosis is the norm in endemic areas like Yucatán, where blood bank testing has served as an important detection route since its implementation in 2013 (following the *NOM-253-SSA1-2012*).

Rural and periurban communities are disproportionately exposed to hematophagous triatomines and, therefore, are at elevated risk of infection and developing CD, while simultaneously facing limited access to healthcare services and diagnostic testing (de Sousa Leite et al., 2024; Ferreira et al., 2025; Jimeno et al., 2021). Improving access to CD screening with point-of-care (POC) diagnostic tests is critical for reaching the most affected, resource-limited populations (Heidt et al., 2020). A range of rapid diagnostic tests (RDTs) has been developed in recent years to support the diagnosis of *T. cruzi* infection. These lateral flow assays offer several operational advantages over conventional serology: they can be distributed and stored at room temperature, utilise low sample volumes generally obtainable via finger-prick capillary blood sampling, and eliminate the need for specialised laboratory infrastructure. Consequently, they can be deployed at the primary care level within local communities, facilitating decentralised testing.

Currently, approximately 25 RDTs are commercially available and evaluated for use in multiple endemic countries (Gabaldón-Figueira et al, 2023). However, RDT performance varies substantially across geographical settings, particularly in terms of sensitivity, highlighting the need for country-specific validation (Perez et al., 2024). In Mexico, independent field-performance data remain limited in Chagas-endemic areas (Ortega-Arroyo et al., 2021; Angheben et al., 2019). Given evidence that conventional serological tests and RDTs may perform less well in Mexico than in other regions (Truyens et al 2021; Ramsey et al 2025; Gamboa-León 2011; Guzmán-Gómez et al 2015), specific evaluation of RDTs is therefore essential to inform their adoption and strengthen diagnostic capacity in the country.

This study aimed to evaluate the diagnostic performance of four commercially available RDTs and one prototype for detecting *T. cruzi* infection in Yucatán, a Chagas-endemic area of Mexico. The results of the RDTs were benchmarked against the diagnostic reference algorithm established by the *NOM*. This approach allowed us to compare the performance of the five RDTs under a common serological reference framework relevant to the Mexican diagnostic context.

## Materials and methods

### Samples

Seventy-nine human sera from Yucatán were analysed, including 49 sera obtained from the Yucatán State Blood Transfusion Centre (*CETS*, Centro Estatal de la Transfusión Sanguínea) and 30 from a previous epidemiological study (*PES*) conducted in Yucatán (González Martínez, 2018) (**Table 1**). The *PES* samples were collected in accordance with ethical approval from the Ethics Board of the Universidad Autónoma de Nuevo León (COBICIS-806/2015/01-FCB-AGM). The *CETS* samples were obtained under authorisation from the Yucatan Health Services SSY/DPCRS/NOR/0899/2025.

**Table 1.**
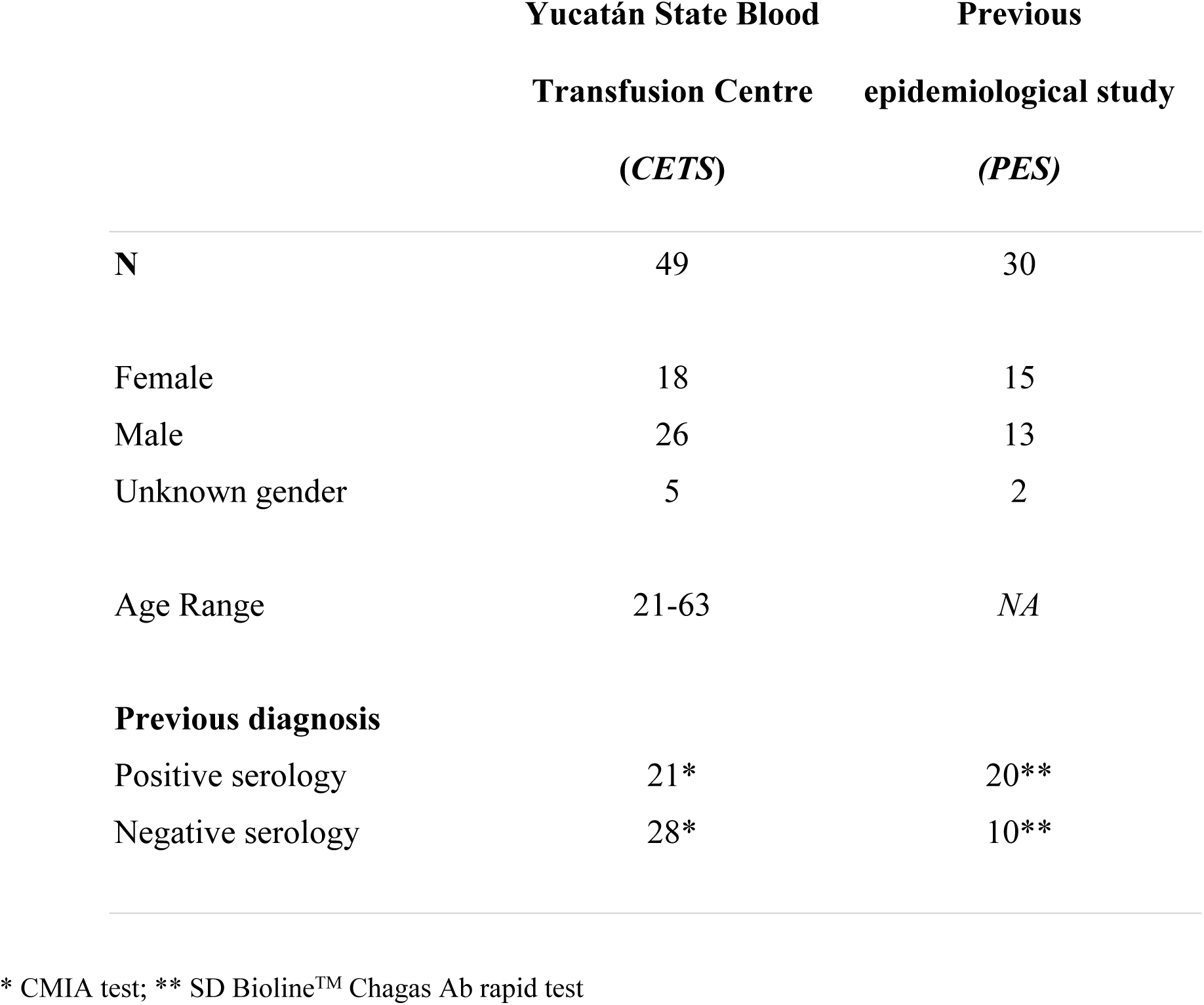
Origin, demographic and previous diagnosis of selected samples.

|  | **Yucatán State Blood**<br>**Transfusion Centre**<br>**(CETS)** | **Previous**<br>**epidemiological study**<br>**(PES)** |
| --- | --- | --- |
| **N** | 49 | 30 |
| Female | 18 | 15 |
| Male | 26 | 13 |
| Unknown gender | 5 | 2 |
| Age Range | 21-63 | *NA* |
| **Previous diagnosis** |  |  |
| Positive serology | 21* | 20** |
| Negative serology | 28* | 10** |

\* CMIA test; \*\* SD Bioline™ Chagas Ab rapid test

Samples obtained from *CETS* were previously classified as positive or negative for *T. cruzi* infection by Chemiluminescent Microparticle Immunoassay (CMIA), while *PES* samples were classified as positive if they had a positive result to the SD Bioline Chagas Ab rapid test (now sold as Bioline^TM^ Chagas Ab by Abbott) in that previous study. All selected samples were anonymised, assigned new random identifiers, and tested in a randomised order. All readers were blinded to the samples’ previous status to minimise potential bias. In order to prevent sample degradation, all sera were kept at-80℃ and aliquots thawed on ice directly before testing.

### Reference serology tests and sample classification

In order to standardise and determine the infection status of all samples, three serological tests (Chagatest ELISA *Lisado*, Chagatest ELISA *Recombinante* v4.0, and Chagatest HAI (product details in Table S1)) were performed. Samples were then serologically re-classified according to the *NOM* diagnostic algorithm. This considers all samples reactive in at least two of the three conventional commercial serological assays to be positive.

### Enzyme-Linked Immunosorbent Assays (ELISAs)

All assays were conducted in the Parasitology Laboratory of the Dr. Hideyo Noguchi Regional Research Centre at the Universidad Autónoma de Yucatán (UADY) according to the manufacturer’s instructions (Chagatest ELISA *Lisado* and Chagatest ELISA *Recombinante* v.4.0; Wiener lab, Rosario, Argentina). Each plate included two positive and three negative controls supplied by the manufacturer and was read at 450 nm. Each assay run was considered valid only when all manufacturer-specified control acceptance criteria were fulfilled.

### Hemagglutination inhibition assay (HAI)

Chagatest HAI was performed according to the manufacturer’s instructions (Chagatest HAI; Wiener lab, Rosario, Argentina). One positive and one negative control were included in each plate. Samples were characterised as positive if a film of cells covered 50% or more of the area at the bottom of the wells, or negative if cells clustered in a small ring at the bottom of the well. Four readers recorded results independently, after which a consensus result was established for each sample by majority decision. In cases where a majority was not reached, the final status was determined via a consensus discussion among the readers.

### Rapid diagnostic tests (RDTs)

A search was conducted within the Mexican National Medical Device Registry (2018– 2025) to identify all Chagas rapid tests with Sanitary Registries issued by the Mexican health authority (*COFEPRIS*, Comisión Federal para la Protección contra Riesgos Sanitarios). Four Chagas rapid tests with Sanitary Registries between 2018-2025 were identified: Chagas Rapid Test (Certum), Bio-Chagas Rapid Test (Mexlab), Bioline^TM^ Chagas Ab (Abbott) and Chagas-Check-1 (Corporinter). An additional test, Chagas Rapid Test (Amunet), was identified as commercially available in Mexico despite the absence of an official Mexican government registration. The manufacturers and suppliers Chagas-Check-1 RDT confirmed that it was not stocked or produced in Mexico at the time of the study; therefore, it was excluded from the project. The other four rapid tests were evaluated, in addition to a prototype under development by Coris: Chagas Sero *K*-SeT. All five products were qualitative membrane-based immunoassays for the detection of anti-*T. cruzi* IgG in whole blood, serum or plasma. All RDTs were used before their expiry date, and tests from each brand were from the same lot (**Table S1**).

All rapid tests were performed immediately after the packaging was opened. Procedures were followed according to the manufacturers’ instructions. For the Bio-Chagas Rapid Test (Mexlab), Chagas Rapid test (Certum), and Chagas Rapid Test (Amunet) cassettes, 25 µL of serum was added to the sample well, followed by two drops of running buffer (approximately 80 µL). In contrast, the Chagas Sero *K*-SeT cassette required 15 µL of serum and 100 µL of running buffer, dispensed into separate chambers. For the Bioline^TM^ Chagas Ab assay, 100 μL of serum was added directly to the cassette; notably, this specific test did not require the use of a running buffer.

Tests were interpreted as positive if a signal on the test (T) line was visible after a 15-minute reaction period. Results were recorded based on the intensity of the test line on a scale of 0 (negative) to 4 (**Fig. S1**). No result was interpreted after 20 minutes of reaction time. The presence of a control (C) line was confirmed for each test. All four readers, blinded to each other’s results, recorded their interpretation independently, and a consensus median score was then calculated for each test. Median scores of ≥ 0.5 were considered positive (tests that were scored positive by at least two readers).

## Statistical analysis

The specificity and sensitivity and Kappa (κ) agreement of *T. cruzi* RDTs was calculated against the *NOM*-based composite reference classification. Sensitivity, specificity and 95% confidence intervals were calculated using the meddecide package (Balci, 2026). Where standard statistical packages failed to compute estimates due to a zero cell count in the 2 x 2 contingency table, sensitivity and specificity were calculated manually as *TP*/(*TP* + *FN*) and *TN*/(*TN* + *FP*), respectively. Exact binomial 95% confidence intervals were computed using the binomial distribution method (Clopper & Pearson, 1934) to appropriately handle boundary values and proportions of 0% or 100%. κ statistics were calculated using the irr package (Gamer and Lemon, 2019) and were classified according to the Landis and Koch scale (Landis and Koch, 1977). The Landis and Koch scale benchmarks inter-rater agreement using six categories ranging from less than 0.00 (poor) to above 0.81 (almost perfect). Diagnostic sensitivities and specificities were compared using the DescTools package to perform Cochran’s Q test, and post hoc pairwise comparisons were conducted using McNemar’s test with Holm correction for multiple testing (Signorell, 2025). Categorical comparisons of correct and incorrect classifications between samples obtained from *CETS* and *PES* were assessed using two-sided Fisher’s exact tests. The Holm-Bonferroni correction was applied to adjust *p-*values for multiple hypothesis testing. The ggplot2 package was used for visualisations (Wickham, 2016). All statistical analysis and plotting were performed in R v4.5.2.

## Results

### Conventional serology

Conventional serological testing was performed on 79 serum samples to determine *T. cruzi* infection status. Using three commercial assays and applying the *NOM* diagnostic algorithm, 36 samples were classified as positive (≥2 reactive tests) and 43 as negative (≤1 reactive test) (**Table 2**). The Chagatest *Lisado* assay did not return any positive results that were discordant with the other two serological assays. However, some discordant positive results were observed with the HAI and *Recombinante* assays. The ELISA *Recombinante* showed greater reactivity than the *Lisado* assay, potentially increasing the likelihood of false-positive results. The HAI assay yielded three isolated reactive results that were classified as negative by the *NOM* algorithm.

**Table 2.**
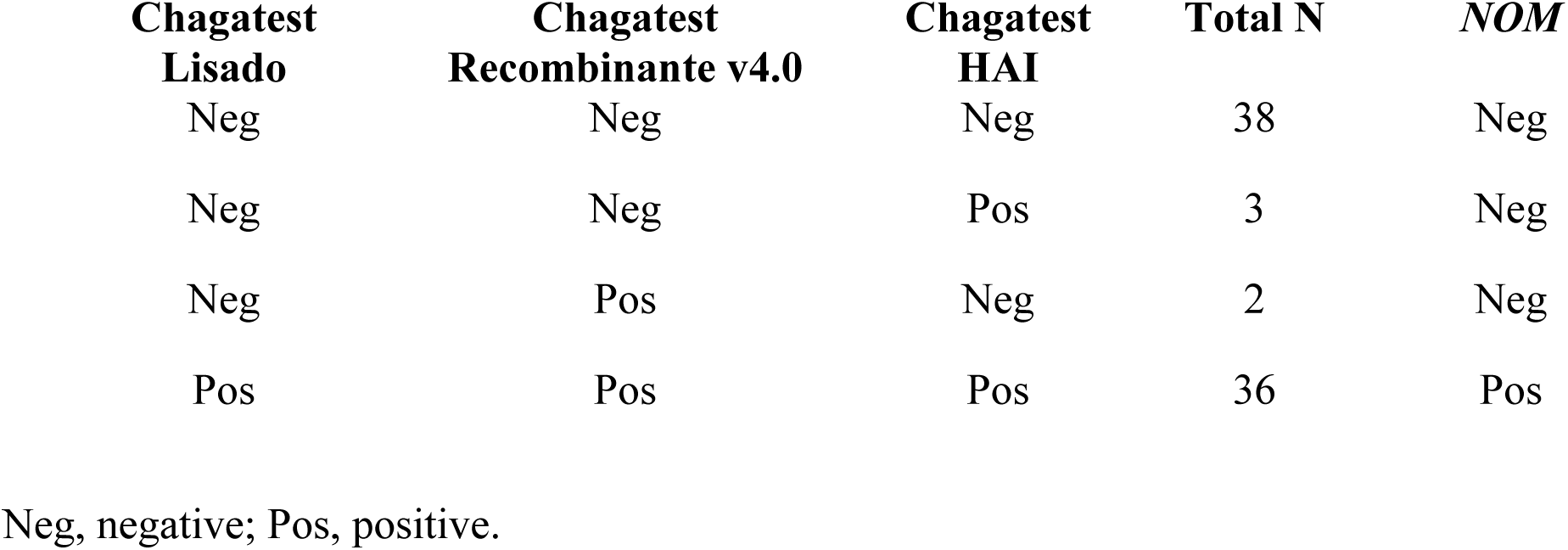
*T. cruzi* infection status according to *NOM* algorithm.

Our sample group included 49 sera that had been previously tested using the CMIA assay routinely employed at *CETS*. Within this *CETS*-tested subgroup (n = 49), the *NOM*-based classification showed an overall agreement of 91.8% with the CMIA result and a κ = 0.83. Only four samples were identified as positive by CMIA but negative by the *NOM* algorithm, with zero false negatives identified. This highlights excellent concordance between the *NOM*-based conventional serology and the blood bank screening testing (**Table 3**).

**Table 3.** Comparison of *NOM*-based serology status to previous CMIA serology status.

| **NOM Status** | **CMIA** | **N (%)** |
| --- | --- | --- |
| Neg | Neg | 28 (57.1) |
| Neg | Pos | 4 (8.2) |
| Pos | Pos | 17 (34.7) |

### Rapid Diagnostic Test Performance

The diagnostic performance of the five RDTs was assessed by comparing their results with the *NOM*-based reference classification. The diagnostic performance of the tests varied significantly according to the RDT. The Bioline^TM^ Chagas Ab test exhibited the highest sensitivity at 97.2%, detecting 35 of 36 positive sera (**Table 4**). The Chagas Sero *K*-SeT prototype also performed well, with a sensitivity of 91.7% (33/36), whereas the Chagas Rapid Test (Amunet) and Bio-Chagas Rapid Test showed significantly lower values of 33.3% and 5.6%, respectively. The Chagas Rapid Test (Certum) failed to detect any positive sera, resulting in diagnostic sensitivity of 0%. In contrast, all evaluated tests demonstrated consistently high specificity, ranging from 93.0% to 100%, indicating a low rate of false positives.

**Table 4.**
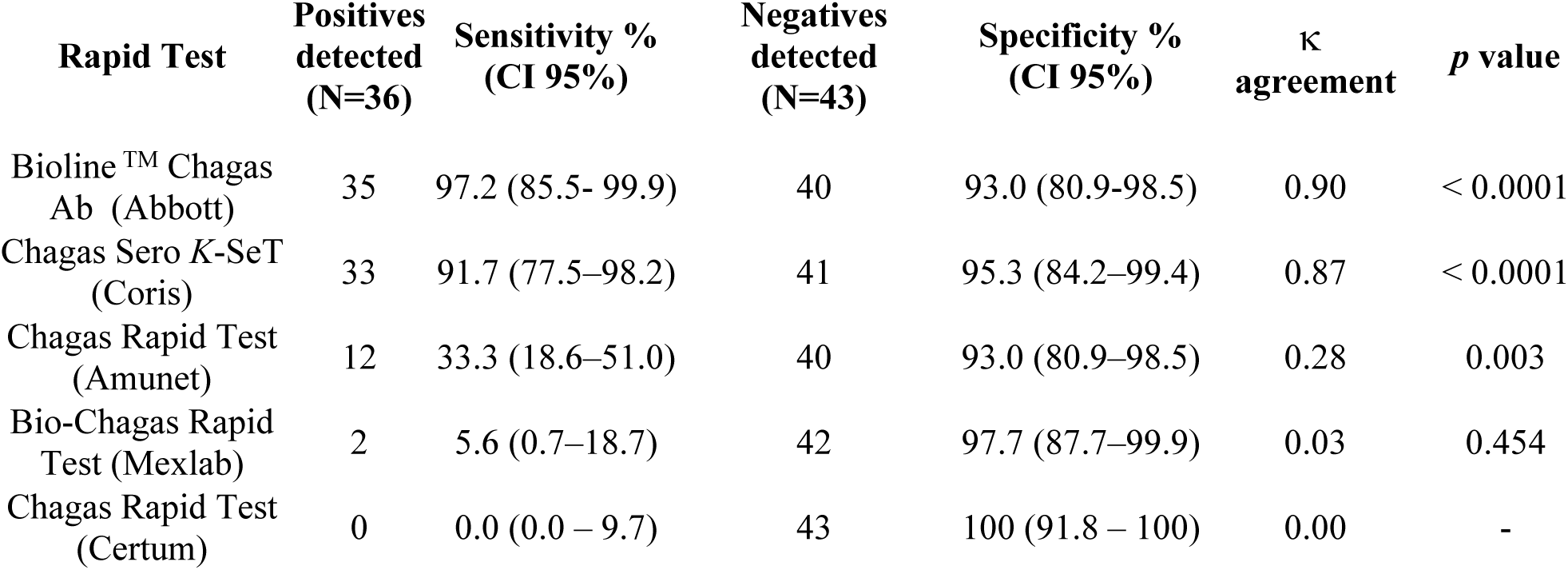
Diagnostic performance of the RDTs evaluated in comparison to the *NOM*-based reference classification.

| Rapid Test | Positives detected (N=36) | Sensitivity % (CI 95%) | Negatives detected (N=43) | Specificity % (CI 95%) | κ agreement | p value |
| --- | --- | --- | --- | --- | --- | --- |
| Bioline™ Chagas Ab (Abbott) | 35 | 97.2 (85.5- 99.9) | 40 | 93.0 (80.9-98.5) | 0.90 | < 0.0001 |
| Chagas Sero K-SeT (Coris) | 33 | 91.7 (77.5–98.2) | 41 | 95.3 (84.2–99.4) | 0.87 | < 0.0001 |
| Chagas Rapid Test (Amunet) | 12 | 33.3 (18.6–51.0) | 40 | 93.0 (80.9–98.5) | 0.28 | 0.003 |
| Bio-Chagas Rapid Test (Mexlab) | 2 | 5.6 (0.7–18.7) | 42 | 97.7 (87.7–99.9) | 0.03 | 0.454 |
| Chagas Rapid Test (Certum) | 0 | 0.0 (0.0 – 9.7) | 43 | 100 (91.8 – 100) | 0.00 | - |

The Bioline^TM^ Chagas Ab and Chagas Sero *K*-SeT tests demonstrated almost perfect agreement with the reference standard (Bioline^TM^ Chagas Ab κ= 0.90, *p* < 0.0001; Chagas Sero *K*-SeT κ = 0.87, *p* < 0.0001), whereas the Chagas Rapid Test (Amunet) test showed fair agreement (κ = 0.28, *p* = 0.002) (**Table 4**). The Bio-Chagas Rapid Test (κ = 0.03, *p* = 0.454) and Chagas Rapid Test (Certum) showed no to slight agreement, reflecting their very low sensitivity for detecting positive samples.

Cochran’s Q test demonstrated a significant difference in sensitivity among the evaluated tests (Q = 107.42, *p* < 0.001), but no significant difference in specificity (Q = 6.182, *p* = 0.186). Post-hoc pairwise McNemar analyses of sensitivity showed that Bioline^TM^ Chagas Ab and Chagas Sero *K*-SeT tests performed comparably (adjusted *p* = 0.317). Both assays demonstrated significantly higher sensitivity than the Chagas Rapid Test (Amunet), Chagas Rapid Test (Certum) and Bio-Chagas Rapid Test (adjusted *p* < 0.001 for all comparisons). The Chagas Rapid Test (Amunet) also demonstrated significantly higher sensitivity than Chagas Rapid Test (Certum) (adjusted *p* < 0.001) and Bio-Chagas Rapid Test (adjusted *p* = 0.002), whereas no significant difference was observed between Chagas Rapid Test (Certum) and Bio-Chagas Rapid Test (adjusted *p* = 0.17).

To address potential bias arising from the pre-selection of *PES* samples using SD Bioline Chagas Ab, we compared the performance of each RDT between the *CETS* and *PES* cohorts. The The Bioline^TM^ Chagas Ab and Chagas Rapid Test (Amunet) RDTs displayed slightly higher sensitivity and specificity in the *PES* samples in comparison to the *CETS* samples. In contrast, the Chagas Sero *K*-SeT displayed slightly higher sensitivity but lower specificity in the *CETS* samples in comparison to the *PES* samples. However, none of these differences in sensitivity or specificity between the two cohorts were statistically significant for any of the five evaluated RDTs (**Table S2**; two-sided Fisher’s exact tests, all Holm-adjusted p = 1.00).

### Intensity scores of the test bands differed between tests

The Bioline^TM^ Chagas Ab and Chagas Sero *K*-SeT tests displayed the full range of intensities (**Fig 1A-B**). The Chagas Rapid Test (Amunet) displayed up to score 2 and the Bio-Chagas Rapid Test score 1 (**Fig 1C**). The Bioline^TM^ Chagas Ab test showed the strongest overall signal (highest median and upper range), while the Chagas Sero *K*-SeT test demonstrated a median signal intensity score of 2. The Chagas Rapid Test (Amunet) demonstrated moderate signal intensity, and the Bio-Chagas Rapid Test yielded predominantly low scores, suggesting weaker antibody capture. The Chagas Rapid Test (Certum) failed to generate measurable intensity in any samples.

**Figure 1.**
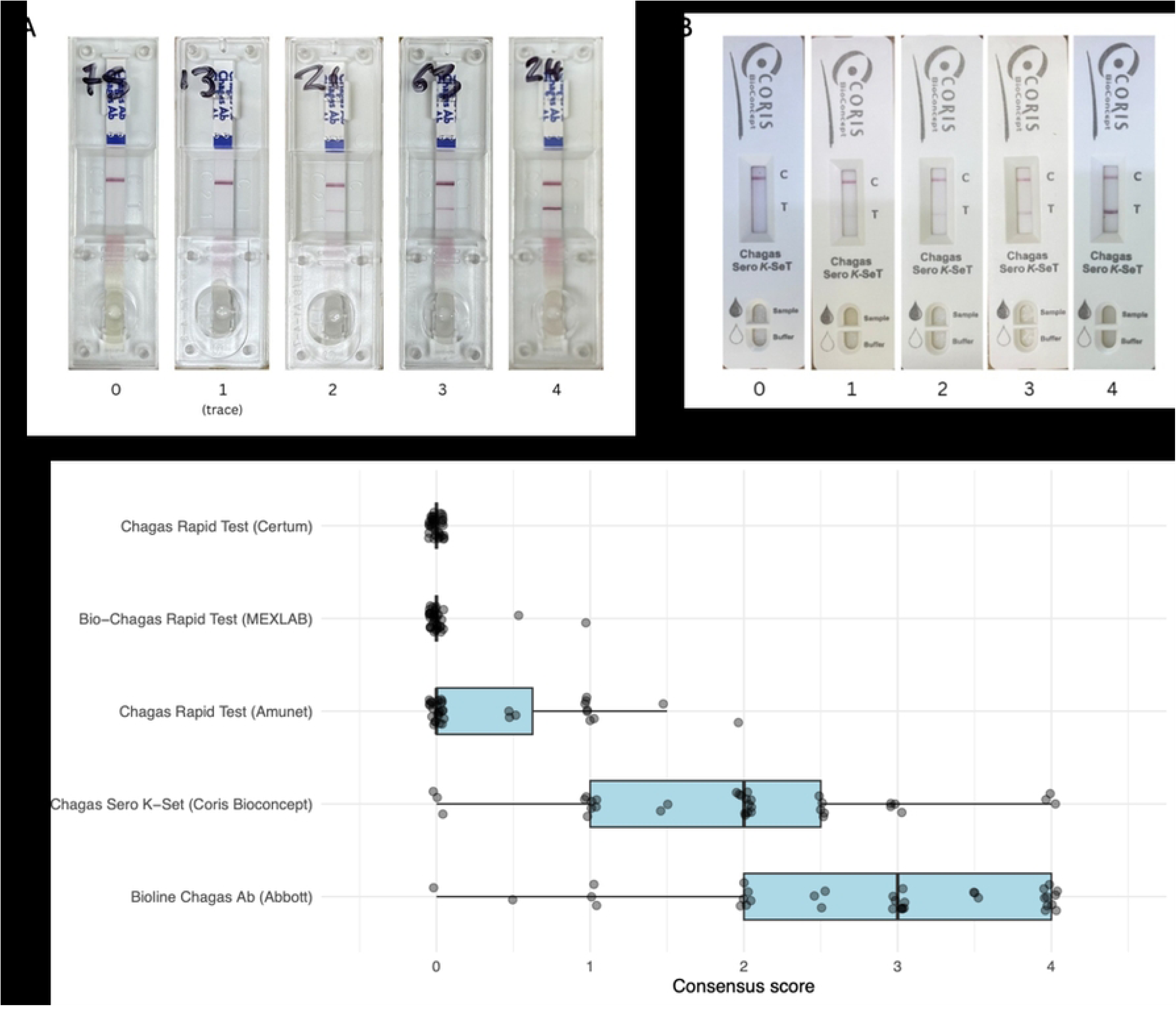
Distribution of RDT line-intensity scores among *NOM*-positive samples. Representative examples of different interpretations of test line intensities for the **(A)** Bioline^TM^ Chagas Ab rapid, and **(B)** Chagas Sero *K*-SeT rapid tests. C, control line; T, test line. **(C)** Box-and-whisker plots display the distribution of consensus RDT scores (0–4) among samples with a positive *NOM*-based classification (N = 36). The vertical line within each box represents the median score, the box indicates the interquartile range (IQR), and whiskers extend to 1.5 × IQR. Individual observations are overlaid as jittered points to illustrate score dispersion.

## Discussion

This study provides an initial assessment of the performance of RDTs for *T. cruzi* infection in samples previously collected in Yucatán, a Chagas-endemic region of Mexico. Firstly, we determined that the *NOM* diagnostic algorithm broadly confirmed previous serological classifications, demonstrating high overall concordance across commercial serology testing platforms. Notably, the combination of Wiener ELISAs (*Lisado* and *Recombinante*) and HAI performed comparably to the CMIA blood bank screening test (91.8% concordance). Importantly, no false negatives were detected by CMIA, supporting its use for blood donor screening.

We then reviewed the performance of five RDTs over samples from the state of Yucatan, and report that Bioline™ Chagas Ab and Chagas Sero *K*-SeT perform comparably to the aforementioned conventional serological tests. The high concordance of these two RDTs with the conventional assays supports their potential use for *T. cruz*i screening, with performance characteristics falling within the WHO desirable ranges for screening tests (Schijman et al., 2024; Porrás et al., 2015). Notably, these two tests utilise distinct antigen targets from the commercial Chagatest *Recombinante* antigens (SAPA, 1, 2, 13, 30 & 36). The concordance of results across assays targeting different antigens provides additional confidence in the validity of the findings. In addition to high specificity and sensitivity, the Bioline^TM^ Chagas Ab and Chagas Sero *K*-SeT RDTs displayed strong signal intensity, which is likely to improve readability and reduce ambiguity in result interpretation compared with tests producing weaker bands.

The Bioline^TM^ Chagas Ab RDT showed a sensitivity of 97.2% and a specificity of 93.0% compared with the *NOM* diagnostic algorithm. These results suggest a similar performance of the Bioline^TM^ Chagas Ab assay compared to studies using sera from Colombia and Brazil, where sensitivities were reported as 86.7% and 95.5%, respectively (Marchiol *et al.,* 2023; Iturra *et al*., 2023). This test uses the 24 kDa calcium-binding protein 1F8/Tc24 and the 300 kDa flagellar protein H49 as antibody targets (Bohorquez *et al*., 2022; da Silveira *et al.,* 2001; Galetović *et al.,* 2011). Both antigens are highly conserved, showing 97% and 94.9% sequence identity across different DTUs, respectively (Maldonando *et al,* 1997; Majeau *et al.,* 2021). Furthermore, these antigens are ubiquitously expressed across all *T. cruzi* life stages (Arnal *et al.,* 2020). Their conservation, together with their expression across all parasite life stages, could explain the test’s consistently high performance across geographically and genetically diverse parasite populations. Furthermore, studies from Argentina have reported that the Bioline^TM^ Chagas Ab test maintains high sensitivity (97.2%) and specificity (97.7%) when used with whole blood samples (Lopez-Albizu *et al*., 2020). The use of whole blood significantly streamlines diagnostic workflows by eliminating laborious sample processing, making it particularly well-suited for primary healthcare settings. However, due to the volume required for this test (100µl), the Bioline^TM^ Chagas Ab would not be ideal for a POC screening test relying on finger-pricked whole blood.

The Chagas Sero *K*-SeT test is currently under development and also shows good overall performance in terms of both sensitivity (91.7%), specificity (95.3%) and signal intensity when performed with human sera from Yucatán. Previous studies in the Chaco region of Argentina have shown that this test performed comparably to an ELISA using the same antibody target, trypomastigote small surface antigen (TSSA), in human and canine samples (Murphy, et al., 2019). The TSSA peptide utilised in this assay has known specificity for TcII, TcV, and TcVI lineages (Bohorquez *et al*., 2022). This design represents an inherent limitation in the context of Mexico, where TcI is the predominant circulating lineage. However, sensitivity remained above 90% in this cohort despite the potential lineage restriction. It is possible that antigenic variation within TcI, partial cross-reactivity, or host immune factors may contribute to the observed performance; however, these possibilities remain speculative. To our knowledge, this is the first reported evaluation of the use of the Chagas Sero *K*-SeT in Mexican populations and, overall, the results justify further evaluating its performance in real-world settings using whole blood.

The study also highlighted variation in the performance of the different RDTs for the detection of *T. cruzi* infection. Of the four RDTs commercially available in Mexico, only the Bioline^TM^ Chagas Ab assay displayed satisfactory diagnostic performance. The poor performance of Bio-Chagas Rapid Test, Chagas Rapid Test (Amunet) and Chagas Rapid Test (Certum) in detecting positive samples is alarming, as they fall far below the standard criteria for diagnostic tools. Our findings suggest that the manufacturer-reported performance of the Chagas Rapid Test (Amunet) and Chagas Rapid Test (Certum) do not adequately reflect their performance in the study region of Yucatán. The observed sensitivities (33.3% and 0%, respectively) represent a profound failure to meet the claimed performance benchmarks (96.7% and 92.9%), raising concerns for clinical use. The performance figures provided by the manufacturers do not clearly specify the geographic origin of the samples or the sample types (serum, plasma, whole blood or synthetic panels) used for validation, which could possibly explain the disparity in results. In the case of the Bio-Chagas Rapid Test, there is no performance data provided by the manufacturer in the product insert. A prior study by Padilla-Vazquez and co-workers reported a total lack of concordance between this test and ELISA-HPX for *T. cruzi* infection (Padilla-Valdez *et al.,* 2025). We shared these results with the manufacturers, and it was recognised that restricted access to characterised sera for test development and optimisation was a common limitation. In this study, collaborating with the Blood Bank of Yucatan (*CETS*) was essential to compile the panel of sera used. This initiative should be continued and replicated in other endemic regions of Mexico, where it could facilitate the establishment of a collection of well-characterised samples. This resource, accessible to both industry and academia, will be crucial for developing new diagnostic tests and evaluating their performance in the Mexican context.

### Study limitations

The selection of samples from a previous epidemiological study may have introduced bias favouring SD Bioline Chagas Ab (now Abbott), as the initial classification of these 30 samples was based on the same assay. This preselection may have led to an overestimation of the test’s sensitivity. Stratification by sample origin showed a sensitivity of 100% in the *PES* samples, compared with 94.1% in samples from *CETS*. However, Fisher’s exact tests did not identify statistically significant differences in sensitivity or specificity between the groups. Given the limited sample size and wide confidence intervals, this lack of statistical significance does not exclude selection bias. Nevertheless, this potential bias does not negate the promising performance observed for Bioline^TM^ Chagas Ab; rather, it highlights the need for further validation in larger, prospectively recruited cohorts that are independent of the index test used for sample selection.

As in many evaluations of diagnostic assays, the study cohort was enriched with previously classified positive cases. This sampling approach should be considered when interpreting the performance estimates and their generalisability to unselected field populations, where disease prevalence and case characteristics may differ. Furthermore, due to the study design, it was not possible to estimate the positive predictive value (PPV) or negative predictive value (NPV) of the RDTs tested, which are also important indicators to consider in control strategies. Larger prospective field studies using whole blood samples in representative populations are therefore needed to more accurately determine diagnostic performance, operational feasibility, and clinical utility of the two best-performing RDTs. Another limitation was the unavailability of paired samples for PCR analysis, which prevented complementary molecular assessment of discordant or ambiguous cases. Recent studies have suggested that conventional serological tests yield limited sensitivity in Mexican cohorts displaying positivity to *T. cruzi* by PCR, possibly due to antigenic diversity in local parasite strains or differences in immune responses of Mexican populations (Ramsey *et al*., 2025; Majeau *et al.,* 2026). Therefore, we cannot rule out the possibility that the sensitivity of the RDTs under study may have been overestimated, as we used only the *NOM*-based reference classification to identify cases of CD.

### Future perspectives and conclusions

Given the promising performance of both tests, Bioline™ Chagas Ab and Chagas Sero K-SeT could be considered candidates for further evaluation as initial POC screening tools for *T. cruzi* infection in Yucatán. These findings represent an initial, Phase II laboratory-based evaluation and support the potential utility of these RDTs for population-level screening in a region recognised as having a high prevalence of CD in Mexico (Ibáñez-Cervantes et al., 2018). However, their performance under routine field conditions remains to be established. Further studies should assess their diagnostic accuracy and operational feasibility in primary care units, including testing by non-specialised healthcare personnel, and evaluate their performance in larger, geographically diverse populations across multiple Mexican states. Multicentre, prospective field validation is essential before these tests can be recommended for routine implementation or incorporated into national diagnostic algorithms.

Furthermore, as the Bioline^TM^ Chagas Ab and Chagas Sero *K*-SeT RDTs target different *T. cruzi* antigens, sequential use warrants evaluation in future studies as a potential POC-based diagnostic algorithm. The second RDT could provide confirmatory evidence based on an independent antigenic target, analogous to the use of two distinct antigens in conventional serological diagnosis under the Mexican *NOM* guidelines and WHO/PAHO recommendations.

In conclusion, we demonstrate for the first time that the Bioline^TM^ Chagas Ab and Chagas Sero *K*-SeT RDTs produced highly promising results with serum samples from Yucatán, performing equally well to the conventional serological tests used for CD diagnosis in Mexico. Furthermore, these results, coupled with the POC attributes of both tests, warrant further investigation into their potential application in real-world scenarios in Yucatán and other regions of Mexico.

## Data Availability

All relevant data supporting the findings of this study are available within the manuscript and its Supporting Information files

## Acknowledgements

We sincerely thank Pascal Mertens and the team at Coris BioConcept for generously providing the Chagas Sero K-SeT rapid diagnostic test prototypes used in this study.

## Author contributions

Conceptualization: J. Alonso-Padilla, B. Bucheton, E. Waleckx

Data curation: R. Heslop, G. Pellegrin, A. Gonzalez-Martínez, C. N. Ibarra-Cerdeña, B. Bucheton, E. Waleckx

Formal analysis: R. Heslop, B. Bucheton

Investigation: R. Heslop, G. Pellegrin, P. P. Martinez-Vega, C. F. Teh-Poot, J. Torres-Montero,

A. Ramos-Ligonio, B. Bucheton, E. Waleckx

Methodology: V. Lejon, C. Guzman-Bracho, J. Alonso-Padilla, B. Bucheton, E. Waleckx

Project administration: B. Bucheton, E. Waleckx

Resources: H. Noh-Pech, K. R. Dzul-Rosado, F. Puerto-Manzano, A. Gonzalez-Martínez, C. N. Ibarra-Cerdeña, C. G. Meza-Ayala, O. Valenzuela-Antelo, J. V. Cruz-Chan, J. A. Ortiz, A. Ramos-Ligonio, B. Bucheton, E. Waleckx

Supervision: B. Bucheton, E. Waleckx

Validation: R. Heslop, G. Pellegrin, B. Bucheton, E. Waleckx

Visualisation: R. Heslop, G. Pellegrin

Writing – Original draft preparation: R. Heslop, G. Pellegrin, J. Alonso-Padilla, B. Bucheton, E. Waleckx

Writing – Review and editing: R. Heslop, G. Pellegrin, P. P. Martinez-Vega, C. F. Teh-Poot, K.

R. Dzul-Rosado, A. Gonzalez-Martínez, C. N. Ibarra-Cerdeña, O. Valenzuela-Antelo, V. Lejon, C. Guzman-Bracho, A. Ramos-Ligonio, J. Alonso-Padilla, B. Bucheton, E. Waleckx.

## Ethics approval and consent to participate

This study was approved by the Research Ethics Committee of the Centro de Investigaciones Regionales Dr. Hideyo Noguchi, Universidad Autónoma de Yucatán (UADY), Mexico (approval number CEI-02-2026).

The study was conducted using 79 previously collected human serum samples from Yucatán, Mexico. Thirty samples originated from a previous epidemiological study approved by the Ethics Board of the Universidad Autónoma de Nuevo León (COBICIS-806/2015/01-FCB-AGM), for which informed consent was obtained from participants. The remaining 49 samples were obtained from the Yucatán State Blood Transfusion Centre (CETS) under authorisation from the Yucatán Health Services (SSY/DPCRS/NOR/0899/2025).

All samples were anonymised and assigned new random identifiers prior to analysis.

## Supplementary figure captions

**Figure S1.** Colour chart used for the interpretation of rapid tests band intensities.

